# Understanding clinician engagement with electronic health records: a mixed-methods study of structured data entry using the Theory of Planned Behaviour

**DOI:** 10.64898/2026.10.08.26365036

**Authors:** Shaun Boustani, Chase Staras, Antonia Rich, Ann Griffin, Andrew J. Wilson

## Abstract

**Background:** Electronic health records (EHRs) are central to NHS clinical practice, and the value of the data they hold depends on clinicians entering structured data. Structured data entry (SDE) is nonetheless used inconsistently, and most evidence comes from US settings documenting barriers rather than the mechanisms interventions must target. This is, to our knowledge, the first study to apply the Theory of Planned Behaviour (TPB) to SDE, to examine SDE behaviour among UK NHS doctors, and to apply corpus linguistics to clinicians’ free-text accounts of EHR use.

**Methods:** A two-part mixed-methods study was conducted at a large NHS teaching hospital in central London using the Epic EHR. Study 1 was a cross-sectional survey of non-consultant grade doctors comprising the System Usability Scale (SUS), a purpose-designed TPB questionnaire, and two open-text questions analysed using corpus linguistics. Study 2 was an Interpretative Phenomenological Analysis (IPA) of semi-structured interviews with six resident doctors.

**Results:** Eighty-three doctors responded; 65 completed the SUS and 49 the TPB questionnaire. Mean SUS score was 62.1 (95% CI 58.3-65.9), below the benchmark of 68 and equivalent to a grade of “D”. Structured features were used markedly less often than narrative notes. TPB constructs explained 56% of the variance in intention (F(3,45) = 18.95, p < .001); attitudes (p = .003) and subjective norms (p = .009) predicted intention independently, whereas perceived behavioural control did not (p = .14). Intention predicted self-reported use of problem lists, event-specific templates and standardised documents, but not narrative notes. Corpus analysis identified time pressure and interface friction as dominant barriers. IPA generated four themes: structured documentation as cognitive relief; as fragile collective infrastructure; as a practice clinicians are expected to perform but not enabled to learn; and as a format that can flatten clinical reasoning.

**Conclusions:** Engagement with SDE is driven principally by attitudes and social norms rather than by perceived control, and the barriers clinicians describe arise from the intersection of interface design, training, team norms and institutional expectation.

Interventions addressing usability alone are unlikely to succeed: sustained change requires team-level normative reinforcement, specialty-relevant training that explains both how and why, and systems that preserve space for clinical narrative.

## Background

Electronic health records (EHRs) have become integral to modern healthcare systems worldwide, transforming how clinical information is captured, shared, and used to support patient care, institutional governance, and health research. In the UK’s National Health Service (NHS), the widespread adoption of systems such as Epic represents one of the most significant structural changes to clinical practice in recent decades, promising improvements in care coordination, patient safety, and data quality [1-3]. When implemented effectively, EHRs can support evidence-based decision-making through clinical decision support tools [4, 5], facilitate communication across multidisciplinary teams [6, 7], and enable patients to engage more actively with their own care [8].

Realising these benefits depends critically on how clinicians engage with EHRs at the point of documentation, and in particular on the quality of the data entered. Structured data entry (SDE) is at the center of this challenge. Where clinical documentation has historically relied on narrative, free-text notes allowing clinicians to record information in their own words and according to their own clinical reasoning, modern EHRs have promoted a shift toward standardised fields, templates, and coding systems designed to produce reusable, interoperable data. In the NHS, where accurate structured coding now underpins both clinical and financial functions, improving SDE quality is a priority for trusts [9, 10]. SDE is also where the needs of clinical reasoning, institutional data collection and patient care can conflict. Templates may flatten complex clinical encounters into rigid categories; structured fields may fail to capture what matters most for individual patient care; and the time and cognitive demands of SDE may sit uncomfortably alongside already pressured clinical workflows [11, 12].

These demands are not trivial. Time-motion and event-log studies show that clinicians spend a substantial and increasing proportion of the working day in the EHR [13, 14], and poor EHR usability has been associated with clinician stress, burnout, and risk to patient safety [15-18]. Navigation and interface design are themselves implicated in documentation error and inefficiency [19], and problem lists, one of the most widely studied structured features, are persistently incomplete across health systems despite their demonstrated value for clinical decision-making [20-22]. Although the present study focuses on doctors, these difficulties are not confined to medicine: nurses, pharmacists and allied health professionals report comparable problems with EHR usability and resistance to documentation change [2, 3, 23]. Understanding why structured documentation is (or is not) taken up therefore matters across the clinical workforce.

Clinicians often recognise the value of structured documentation in principle while finding it difficult to enact in practice [24], suggesting that barriers to SDE cannot be explained by usability or training alone. How clinicians think about SDE, what their colleagues and institutions expect of them, and how documentation demands fit (or do not fit) the realities of clinical work all shape engagement in ways that technical fixes are unlikely to address [15, 23, 25]. Understanding SDE behaviour therefore requires a framework that attends to the psychological, normative, and experiential dimensions of clinical documentation practice.

The Theory of Planned Behaviour (TPB) provides such a framework [26]. The TPB proposes that behaviour is most proximally determined by behavioural intention, which is in turn predicted by attitudes toward the behaviour, subjective norms (perceived social pressure), and perceived behavioural control (PBC). The evidence base for the model is substantial: meta-analytic work across behavioural domains shows that TPB constructs typically account for around 39% of the variance in intention and 27% of the variance in behaviour [27], and reviews of health-related applications confirm its predictive utility in healthcare contexts specifically [28]. The model has been applied successfully to healthcare professionals’ own behaviour, including doctors’ intentions to engage in professional behaviours such as raising patient safety concerns and reflective practice [29], and healthcare professionals’ intentions to use clinical guidelines [30]. Established methodological guidance exists for constructing domain-specific TPB questionnaires [31, 32]. Applying the TPB to SDE therefore offers a validated, replicable means of identifying which psychological levers an intervention would need to pull, and of comparing findings directly with the wider implementation literature. Figure 1 summarises the model as applied in this study.

**Figure 1.**
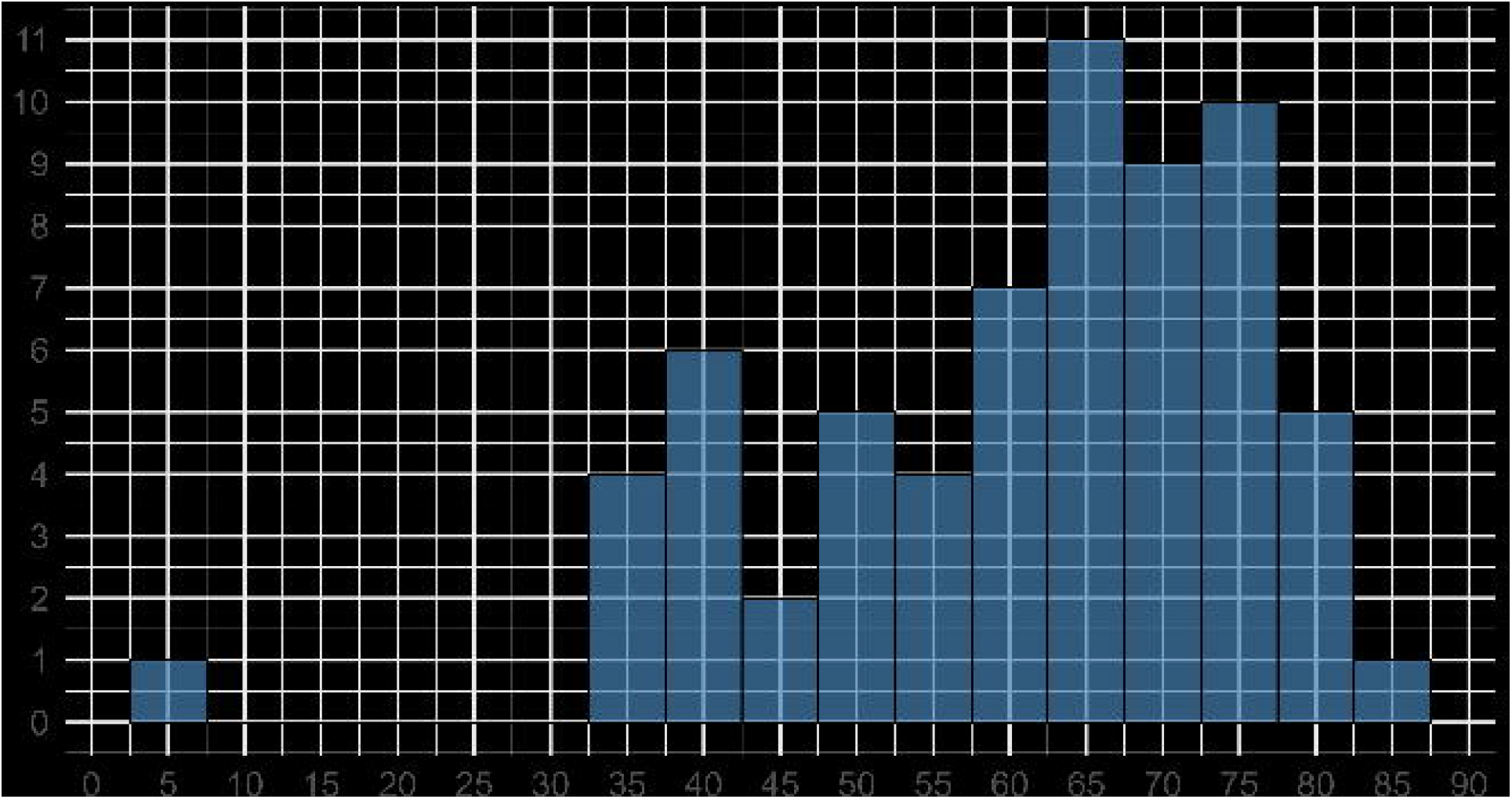
The Theory of Planned Behaviour as applied to structured data entry. Attitudes, subjective norms and perceived behavioural control predict behavioural intention, which in turn predicts behaviour; perceived behavioural control is also theorised to influence behaviour directly. Adapted from Ajzen [26].

Most existing research on SDE has been conducted in US settings and has tended to document barriers or to test discrete interventions without examining the underlying mechanisms that would need to be targeted for those interventions to succeed [33, 34]. US health systems operate under organisational and financial conditions that differ substantially from those of the NHS, particularly in the role that coded documentation plays in reimbursement, so the transferability of those findings to a UK setting cannot be assumed. Addressing this gap requires understanding both what clinicians think and feel about SDE, and what social and institutional expectations bear on their documentation behaviour, and how the practice is experienced and made meaningful in the realities of everyday clinical work.

### The current study

We used a different method for each of these questions. Clinicians’ attitudes toward structured documentation, the expectations of colleagues and institutions, and their sense of control over documentation are the constructs measured by the TPB [26]. The experiential dimension, detailing how SDE is lived, negotiated, and made meaningful in everyday clinical work, requires a qualitative approach capable of attending to meaning-making and situated practice; for this we employed Interpretative Phenomenological Analysis (IPA) [35].

This study therefore adopts a mixed-methods design to assess current perceptions of the EHR system in use at University College London Hospitals NHS Foundation Trust (UCLH), a large teaching hospital in central London, and the predictors of intention to enter structured data into that system. Study 1 used a theory-driven quantitative survey to examine EHR usability and the predictors of non-consultant grade doctors’ intentions to engage in SDE. Study 2 employed IPA to explore how SDE is experienced and made meaningful in everyday clinical practice. This study reports the first stage of a broader programme of work aimed at improving EHR usability and data quality at the Trust; findings will inform the subsequent design and evaluation of clinician-facing interventions, which will be reported separately.

The two studies adopt different epistemological orientations. Study 1 is grounded in a positivist framework aimed at modelling relationships between beliefs, norms, and intentions. Study 2 is interpretative and idiographic, concerned with how resident doctors experience and make sense of SDE in context. We do not treat the qualitative findings as tests of the quantitative model. Instead, the studies are positioned as complementary: the first addresses what predicts intention at a population level, the second examines how the practice is lived, negotiated, and understood in everyday work.

The study had the following objectives:

1. To assess non-consultant grade doctors’ current perceptions of the EHR at UCLH.
2. To understand the baseline attitudes, subjective norms, perceived behavioural control, and intentions of doctors to engage in EHR structured data entry.
3. To evaluate perceived barriers to EHR usage.
4. To explore the lived experience of structured data entry through in-depth qualitative interviews.

## Methods

### Study design

A convergent mixed-methods design was used. Study 1 was a cross-sectional survey containing validated quantitative scales and open-ended questions; Study 2 was a qualitative interview study analysed using IPA. Both studies were conducted at UCLH, a large NHS teaching hospital in central London that uses the Epic EHR. Reporting follows the STROBE statement for Study 1 and the COREQ checklist for Study 2 (Additional Files 3 and 4).

### Study 1: Survey

#### Participants and recruitment

Recruitment was managed by UCLH, with the survey administered trust-wide through Trust mailing lists and in-person teaching events, using Qualtrics. The survey was open from 21 January to 8 September 2025. All non-consultant grade doctors employed by the Trust were eligible to participate. This included resident doctors (FY1–2, core and specialty trainees, and GP specialty trainees), Locally Employed (Trust) doctors, who are non-training grade clinicians employed directly by the Trust, and Specialist, Associate Specialist and Specialty (SAS) doctors, who are clinicians with significant clinical experience who have completed or partially completed specialty training but do not hold a consultant post. Consultant-grade doctors were not eligible: consultants’ documentation responsibilities and patterns of EHR use differ substantively from those of non-consultant grade doctors, who undertake the majority of routine inpatient documentation at the Trust, and who were the intended users of the interventions this programme of work is designed to inform.

Because the survey was distributed both through Trust-wide mailing lists and at in-person teaching events, the number of doctors who received or saw an invitation could not be determined, and a response rate therefore cannot be calculated. The Trust employs approximately 1,000 non-consultant grade doctors, but this is an estimate of the eligible workforce rather than of the number invited, and it is not used here as a denominator. The implications of this for interpreting the sample are considered in the Limitations. The survey received 83 responses. Of these, 65 completed the System Usability Scale (SUS) and 49 completed the TPB questionnaire. Respondent characteristics are shown in Table 1. Age and gender were not collected, in order to minimise the risk of deductive disclosure in a single-site sample.

**Table 1.**
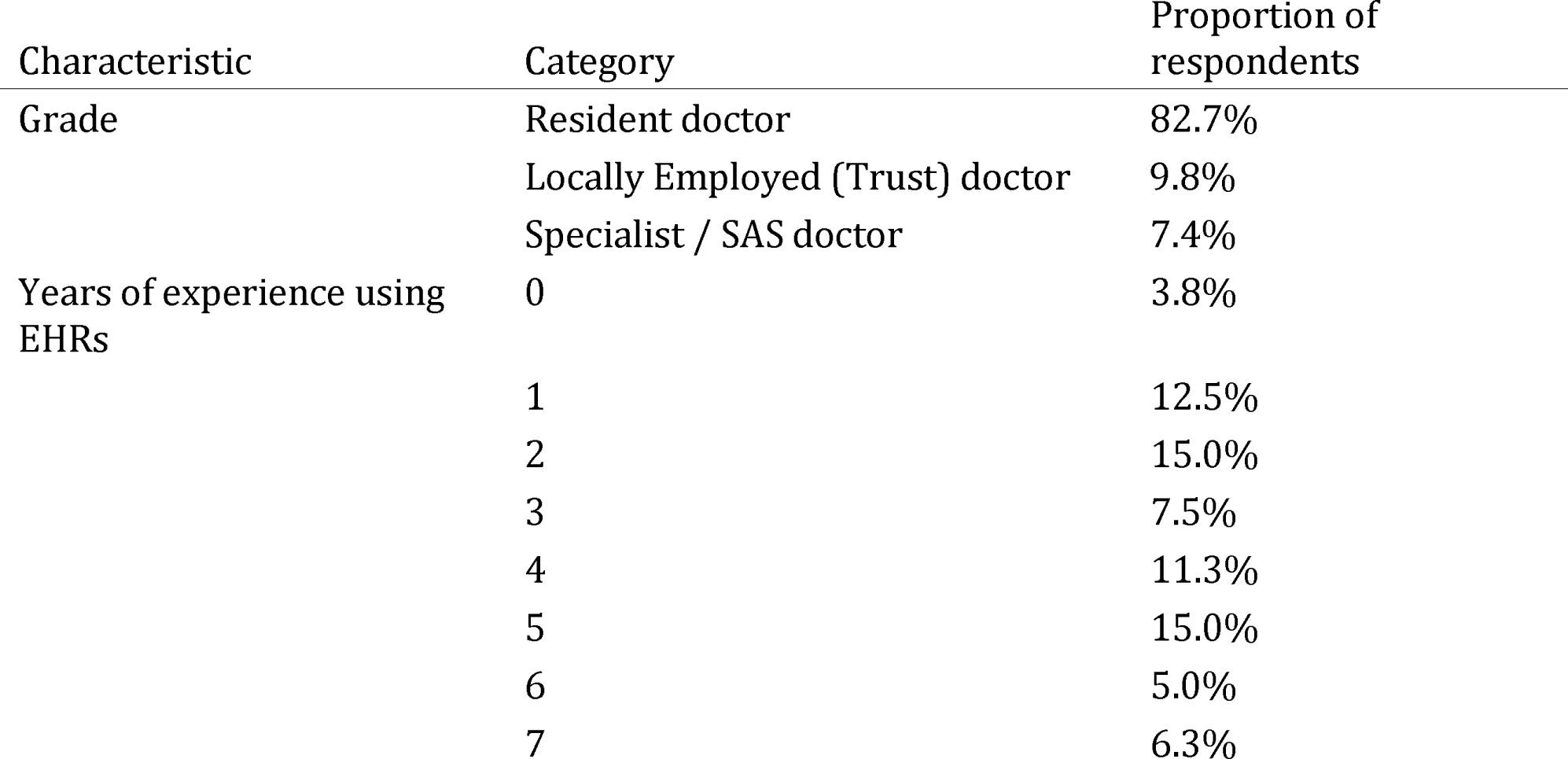
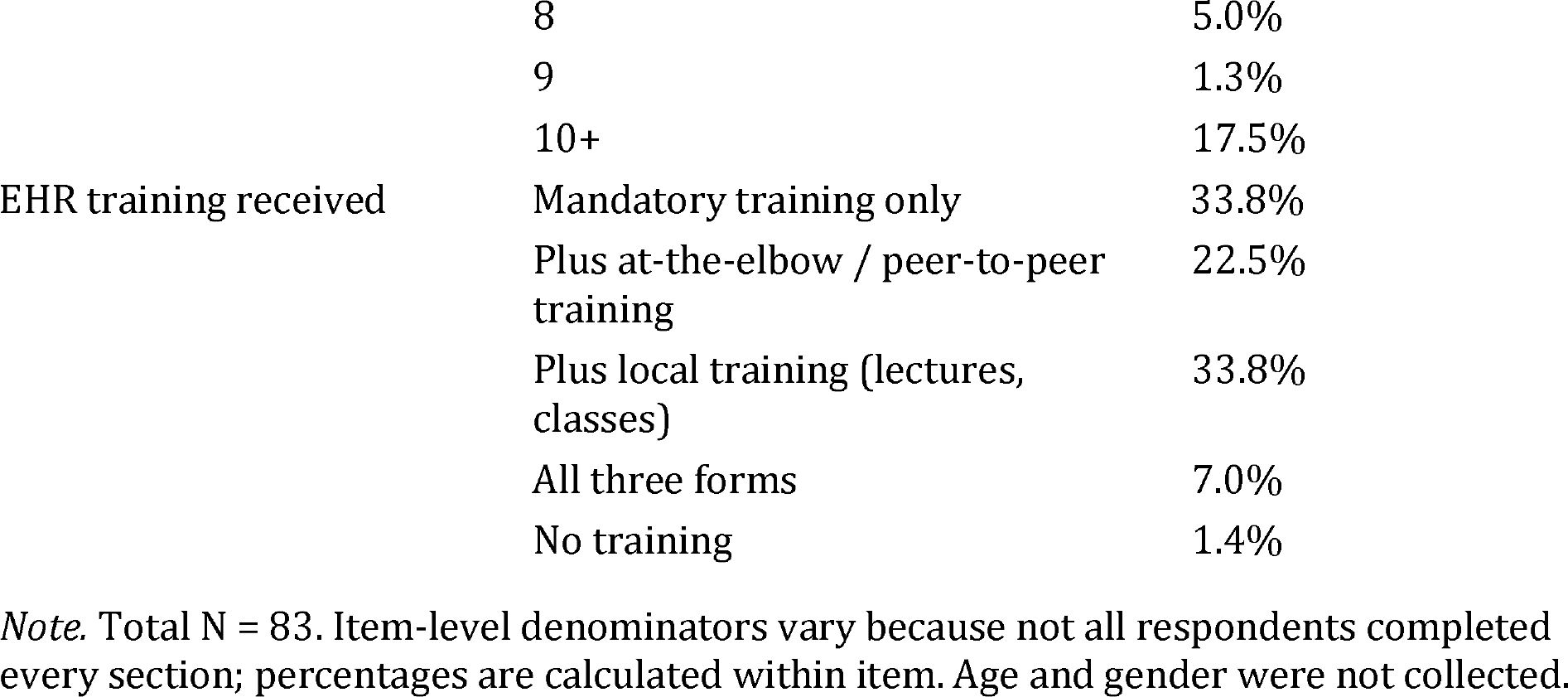
Respondent characteristics, Study 1 survey.

#### Survey design and measures

The survey was developed with input from clinical and informatics colleagues and refined following informal piloting before distribution. The final instrument assessed the EHR in use at the Trust across two principal components, alongside items on training and on the self-reported frequency of use of specific EHR features. The full instrument is provided in Additional File 1.

##### Theory of Planned Behaviour questionnaire

The TPB questionnaire was developed in accordance with established guidance for constructing TPB measures [31, 32]. The target behaviour was defined for participants at the outset as entering formatted data using problem lists and event-specific templates (such as procedure notes, consultation notes, and operation notes) rather than default options such as progress notes or patient narratives, and was specified throughout as “entering structured data in the Epic EHR system during or immediately after every patient consultation during my shifts”, giving a consistent action, target, context and time across all items in line with the principle of compatibility [31]. Definitions of problem lists and event-specific templates were displayed before the section began. All TPB items were rated on seven-point bipolar scales. Intention was measured with three items (for example, “I intend to enter structured data in the Epic EHR system during or immediately after every patient consultation during my shifts”, rated extremely unlikely to extremely likely). Attitudes were measured with four semantic differential items anchored harmful-beneficial, pleasant-unpleasant, good-bad and worthless-valuable. Subjective norms were measured with five items comprising two injunctive items (for example, “It is expected of me that I enter structured data…”) and three descriptive items (for example, “Most colleagues who are important at work enter structured data…”). Perceived behavioural control was measured with four items, two addressing capacity (“For me to enter structured data… would be very difficult/very easy”; “If I wanted to, I could enter structured data…”) and two addressing controllability (“How much control do you believe you have over entering structured data…?”; “It is mostly up to me whether or not I enter structured data…”). Two items, one from each pair, were removed during scale refinement because they lowered internal consistency, leaving a two-item composite comprising one capacity and one controllability item. Items with reversed anchors were reverse scored so that higher scores indicated a more favourable orientation in every case, and item order was randomised. A worked practice item was presented before the section to familiarise respondents with the seven-point format. The full instrument is reproduced in Additional File 1.

##### System Usability Scale

Current EHR usability was assessed using the System Usability Scale (SUS) [36]. The SUS is a standardised, validated and widely used instrument consisting of 10 statements rated on a five-point Likert scale ranging from “Strongly disagree” to “Strongly agree”, with items alternating between positive and negative phrasing. Raw item scores are adjusted, summed and multiplied by 2.5 to produce an overall score ranging from 0 to 100, interpreted as a composite measure of perceived usability, with higher scores indicating better usability. The only adaptation to the standard instrument was the substitution of “this EHR system” for “this system” in each item, with respondents instructed to rate the EHR system in use at UCLH (Epic); no items were added, removed or otherwise reworded, so scores remain comparable with published SUS benchmarks. The ten items as administered are reproduced in Additional File 1. Scores were interpreted against the established benchmark of 68, which represents average usability across systems and is also the reported average for digital health applications [37], and against the Sauro-Lewis Curved Grading Scale, which converts numeric scores to letter grades [38]; adjective ratings were interpreted following Bangor and colleagues [39].

##### Open-text items

Participants were asked two open-ended questions: what the biggest barriers were to using Epic to input structured data, and what suggestions they had for improving the system.

#### Analysis

Composite scores for each TPB construct were calculated as the mean of the available constituent items after reverse scoring. Internal consistency was assessed using Cronbach’s alpha, with a threshold of acceptability of α = 0.60 set a priori [40]; scales were refined by removing items that lowered alpha, as described above for perceived behavioural control. Relationships among TPB constructs were examined using Pearson correlations, and a multiple linear regression was used to predict behavioural intention from attitudes, subjective norms and perceived behavioural control. Because the subjective norms scale combined injunctive and descriptive items, a sensitivity analysis entering the two as separate predictors was also run; it did not improve on the combined model, which is therefore reported. A further series of simple linear regressions examined intention as a predictor of the self-reported frequency of using each documentation method. Regression models used listwise deletion, which accounts for the small differences in degrees of freedom between models. Analyses were conducted in R version 4.3.2 using the psych package for reliability analysis. With 49 complete TPB responses, the multiple regression was powered to detect a large effect (f^2^ = 0.35) with three predictors at α = .05 with approximately 80% power, but was underpowered to detect small independent contributions; this is considered further in the Discussion.

Open-text responses were analysed using corpus linguistics in LancsBox X. Corpus linguistics is a quantitative approach to text analysis that identifies which words occur most frequently and determines whether their frequency is significantly higher than would be expected by chance, by comparison with a reference corpus; here the Spoken BNC2014 was used as the reference corpus [41, 42]. We report only keywords reaching a Simple Maths threshold greater than 10 and a log-likelihood value above 6.63, indicating that their frequency was statistically greater than baseline [43]. Because the dataset was small, we examined only words appearing more than 10 times or in more than 10% of comments. Concordance analyses, in which the immediate textual context of each keyword is reviewed, were conducted for all reported keywords [44].

#### Ethics

Ethical approval was granted by the UCL Research Ethics Committee before the study commenced (approval ID 005). All participants were provided with an information sheet and gave informed consent electronically before beginning the survey. Participants were informed that the survey would take approximately 10 minutes, that responses were anonymous and no personally identifying information would be collected, that participation was voluntary, and that they could withdraw at any point before submitting. The same approval covered Study 2.

#### Study 2: Interpretative Phenomenological Analysis

While the survey identified key predictors of intention, it raised several questions that could not be addressed through quantitative work: what makes SDE experienced as more cumbersome than unstructured documentation; how resident doctors understand and evaluate available training; how social norms around SDE operate in practice; and which contextual features of clinical work shape perceived usefulness and uptake. These questions concern meaning, sense-making and situated practice rather than variable-level relationships, and therefore required a qualitative approach. At the time of data collection, the haematology department, where most participants worked, had already gone live with SDE templates for ward round notes, meaning that participants were drawing on direct lived experience of structured documentation rather than anticipated use.

#### Design

To explore resident doctors’ lived experience of SDE, we conducted a qualitative study using IPA [35]. IPA was selected because it is designed to examine how individuals make sense of complex, contextually embedded practices, and is appropriate for small, purposively sampled datasets where depth of analysis is prioritised over representativeness [45]. IPA has been used extensively in healthcare research to examine health professionals’ experiences of demanding or disruptive aspects of practice, including burnout [46] and multidisciplinary collaboration [47], and the adjustment of early-career doctors to new professional roles [48]. The introduction of structured documentation represents a comparable transition, involving changes to documentation practices, to the division of clinical labour, and to the way clinical reasoning is recorded and made visible to others.

#### Participants and recruitment

Participants were recruited using purposive sampling from resident doctors employed within the Trust who used Epic as part of their routine clinical practice. Eligibility criteria required participants to be resident doctors, working clinically within the Trust at the time of the study, and regular users of Epic for patient documentation; these criteria defined a deliberately homogeneous group for idiographic analysis. An invitation to participate was circulated via departmental mailing lists sent by the Trust to the same population approached for Study 1. Distribution was managed by the Trust as gatekeeper, so the number of doctors who received the invitation is not known. Doctors who wished to take part responded directly and provided digital consent online, following which they were contacted by SB and CS to arrange an interview.

Six resident doctors consented to take part. Five were based in haematology and one in neurosurgery. In line with IPA guidance, all participants shared the same career stage, organisational context and EHR system, providing a relatively homogeneous sample for idiographic analysis [35]. Participant characteristics are provided in Table 2.

**Table 2.** Study 2 participant characteristics.

| Pseudonym | Specialty | Grade | Time in present role |
| --- | --- | --- | --- |
| Amanda | Haematology | Registrar | 5 years |
| Jay | Haematology | MST7 | 7 years |
| Martin | Neurosurgery | Junior neurosurgical resident | 4 months |
| Ryan | Haematology | Registrar | 8 months |
| Vincent | Haematology | ST2 | 1 year |
| Yusef | Haematology | ST5 | 4 months |

*Note.* All names are pseudonyms.

#### Data collection

Data were collected through semi-structured, one-to-one online interviews conducted using Microsoft Teams. The interview guide (Additional File 2) was developed to support a phenomenological understanding of attitudes (for example, “What do you think about using structured fields?”), social norms (for example, “Do your peers and senior colleagues use structured data entry differently to you?”), and perceived behavioural control (for example, “Do you feel that you can use the EHR in the way we have been discussing?”) with regard to SDE. These TPB domains served as sensitising concepts rather than analytic categories: questions were intentionally open and experience-focused, and the guide was used flexibly to support participants’ own accounts. Prompts were used to encourage elaboration, reflection and concrete examples, in line with IPA’s emphasis on meaning-making and experiential depth [35].

Interviews were conducted by SB (PhD, Senior Research Fellow, male) and CS (PhD, Research Fellow, male), both of whom have prior experience in qualitative interviewing; SB brought expertise in the TPB and CS familiarity with IPA. Neither interviewer was known to any participant before the study, and neither held a clinical or supervisory role at the Trust. Participants were told that the researchers were independent university-based researchers at UCL. Only the participant and the interviewer were present at each interview, and no repeat interviews were conducted. Interviews took place between 5 and 27 November 2025 and lasted an average of 29 minutes (range 23–33 minutes). All interviews were audio and video recorded with participants’ written and verbal consent, transcribed using Microsoft Teams transcription software, and checked against the recordings for accuracy. Brief handwritten notes were made during interviews as an aide-memoire but were not used in analysis. Transcripts were not returned to participants for comment. Identifying information was removed to preserve anonymity, and all data were stored securely on institutional OneDrive.

#### Data analysis

Data were analysed by CS using IPA following the steps outlined by Smith and colleagues [35], with NVivo used to manage the data. Each transcript was first read and re-read to develop familiarity with the participant’s narrative. Initial exploratory coding attended to descriptive content, linguistic features and conceptual reflections. Personal Experiential Themes (PETs) were developed for each participant before PETs were examined across participants to develop Group Experiential Themes (GETs) capturing patterns of convergence and divergence across the corpus. Analysis was not structured around TPB constructs; themes were developed inductively from participants’ accounts, allowing experiential concerns to emerge that cut across or exceeded the domains informing the interview guide.

Throughout analysis, CS reflected on their role in making sense of participants’ sense-making of their own experiences, consistent with IPA’s double hermeneutic [35]. Analytic decisions were documented and discussed between authors to ensure that interpretations remained grounded in participants’ experiences. Although GETs were developed across participants, the analytic priority remained the preservation of the idiographic integrity of each account. Quotes were selected to evidence each theme and to represent convergence and divergence across the sample. Participants were not asked to provide feedback on the findings.

## Results

### Study 1: Survey

#### Respondent characteristics

Eighty-three doctors responded to the survey. The majority (82.7%) were resident doctors; the next most frequent roles were Locally Employed doctors (9.8%) and Specialist doctors (7.4%). Respondents reported between 0 and 10 or more years of experience using EHRs, with 10 or more years the single most common response (17.5%) and the majority (61.4%) reporting four or more years. Respondent characteristics are summarised in Table 1. Item-level denominators vary because not all respondents completed every section.

#### EHR training

Almost all respondents (98.6%) reported having had some form of EHR training; one respondent reported none. A third (33.8%) had completed mandatory training only, 22.5% had additionally received at-the-elbow or peer-to-peer training, 33.8% had received additional local training such as lectures or classes, and 7.0% had received all three forms.

Most respondents reported that their training had at least adequately prepared them for using Epic: 62.9% responded “Somewhat” and 10.0% “To a great extent”, while 20.0% responded “Very little” and 7.1% “Not at all”.

Respondents were prompted to leave an optional open-text comment elaborating on their answer, and 26 did so. Content analysis of these comments identified two themes. Fifteen respondents expressed a preference for active training, that is, training that was hands-on, delivered on the job, or non-didactic. Nine commented that their training either lacked clinical or specialty relevance, or was too generic or basic.

#### Frequency of EHR feature use

Participants were asked how frequently they used each of a set of EHR features (Table 3). The feature most often reported as used “always” or “most of the time” was the default narrative (progress notes; 72.9% combined). Structured features were used considerably less often: only 17.4% of respondents used problem lists always or most of the time, compared with 46.3% for event-specific templates and 48.5% for departmental notes.

**Table 3.** Self-reported frequency of use of EHR features (%).

| Frequency | Problem lists | Default narrative (progress notes) | Event-specific templates | Departmental notes | Other features |
| --- | --- | --- | --- | --- | --- |
| Always | 8.7 | 42.9 | 27.5 | 27.1 | 14.0 |
| Most of the time | 8.7 | 30.0 | 18.8 | 21.4 | 16.0 |
| About half the time | 14.5 | 7.1 | 8.7 | 12.9 | 6.0 |
| Sometimes | 39.1 | 5.7 | 21.7 | 18.6 | 24.0 |
| Never | 29.0 | 14.3 | 23.2 | 20.0 | 40.0 |

Participants who reported using other features were prompted to describe them; 17 did so. Of these, nine (53%) mentioned smart phrases, five (29%) mentioned other types of template (for example, self-created templates or letter templates), one mentioned specialty-specific features (anaesthetic documents), and two mentioned research-related features.

#### EHR usability

The mean SUS score for the sample was 62.1 (95% CI 58.3–65.9). Scores ranged from 2.5 to 85, with a median of 65 and a mode of 67.5 (Figure 2). A SUS score of 68 is the established benchmark for average usability and is also the reported average for digital health applications [37]; the observed mean therefore falls below benchmark and corresponds to a grade of “D” on the Sauro-Lewis Curved Grading Scale [38]. This indicates that users are experiencing meaningful usability difficulties and that there is substantial room for improvement.

**Figure 2.**
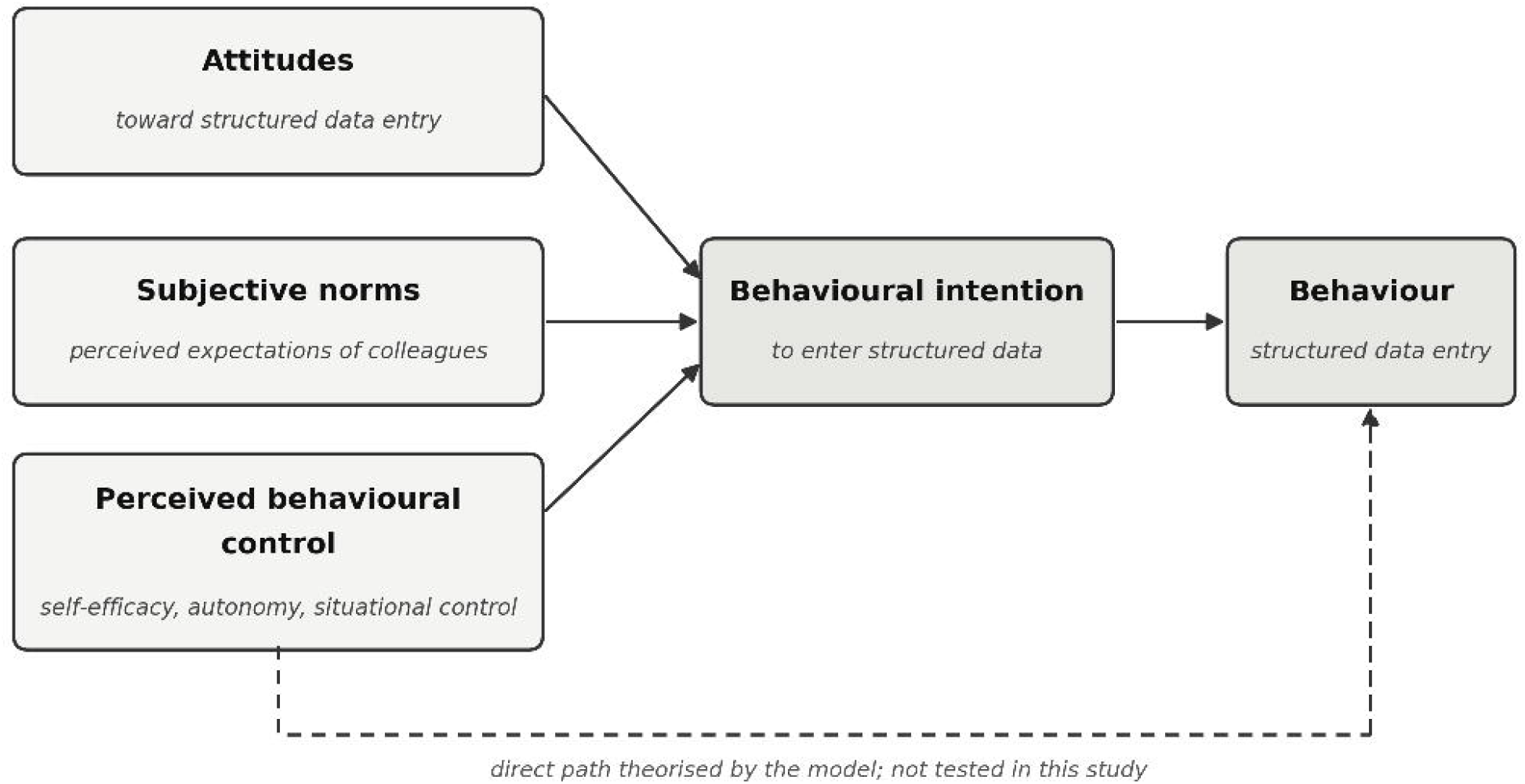
Distribution of System Usability Scale scores (n = 65). The dashed line indicates the established benchmark score of 68 for average usability. Mean = 62.1 (95% CI 58.3– 65.9); median = 65.

#### Theory of Planned Behaviour analysis

##### Reliability

Internal consistency was acceptable for all scales: intention (3 items) α = 0.88; attitudes (4 items) α = 0.81; subjective norms (5 items, combining injunctive and descriptive items) α = 0.83; and perceived behavioural control (2 items retained of 4 administered) α = 0.73. All exceeded the a priori threshold of 0.60 [40].

##### Descriptive statistics

Mean composite scores on the seven-point scales were modest and clustered just above the scale midpoint: intention M = 4.40 (SD = 1.61), attitudes M = 4.55 (SD = 1.25), subjective norms M = 4.10 (SD = 1.38), and perceived behavioural control M = 4.01 (SD = 1.26).

##### Relationships among constructs

Intention correlated moderately with attitudes (r = 0.66), subjective norms (r = 0.57) and perceived behavioural control (r = 0.57), indicating that while the constructs share common variance, each also captures distinct aspects of the decision-making process.

A multiple regression predicting behavioural intention from attitudes, subjective norms and perceived behavioural control produced a significant overall model, F(3, 45) = 18.95, p < .001, R^2^ = .56. Examination of individual predictors indicated that attitudes (B = 0.53, 95% CI 0.19 to 0.86, t = 3.17, p = .003) and subjective norms (B = 0.36, 95% CI 0.10 to 0.63, t = 2.75, p = .009) were significant independent predictors of intention, whereas perceived behavioural control was not (B = 0.25, 95% CI −0.08 to 0.58, t = 1.50, p = .14). Together these predictors explained 56% of the variance in intention, underscoring the central role of evaluative and normative beliefs in shaping intentions toward SDE.

##### Prediction of behavioural outcomes

A series of regression analyses examined the effect of behavioural intention on the self-reported frequency of using different documentation tools. Intention was a significant predictor of the frequency of entering problem lists (B = 0.24, 95% CI 0.01 to 0.48, t(46) = 2.12, p = .039), event-specific note templates (B = 0.29, 95% CI 0.02 to 0.56, t(46) = 2.17, p = .035), and standardised documents (B = 0.33, 95% CI 0.09 to 0.57, t(47) = 2.71, p = .009), but not narrative notes (B = −0.04, 95% CI −0.28 to 0.20, t(47) = −0.32, p = .752). Higher intention to engage in SDE was therefore associated with increased use of features typically considered structured, but not with the use of unstructured narrative notes.

#### Open-text responses

Forty-four respondents answered the question on barriers and 37 the question on suggested improvements.

##### Barriers

The most frequently reported barriers to using Epic for SDE related to time constraints and technical difficulty (Table 4). Nearly half of all responses (47.7%) cited time, indicating that data entry is perceived as excessively time-consuming. Problem lists were mentioned in 18.2% of responses, and “data” and “structured” in 15.9% and 13.6% respectively. Concordance analysis revealed that respondents using these terms were commenting on the need to fill in, code or maintain problem lists manually, the clunkiness of the user interface, inconsistency of use across colleagues, and inaccuracies in the data. Comments on the quality and quantity of “information” and on lag or clunkiness of the Epic “system” each appeared in around 14% of responses. Analysis of responses mentioning “notes” and “note” (18.2% and 11.4%) indicated that respondents often favoured less structured forms of documentation, such as ward, clinical and progress notes, which they described as easier to use, more time-efficient, and occasionally more detailed than the structured options the system provides.

**Table 4.** Keywords from corpus analysis of open-text responses on barriers to EHR use.

| Keyword | Simple Maths | Log-likelihood | Frequency | Range | Range % |
| --- | --- | --- | --- | --- | --- |
| time | 11.86 | 80.43 | 25 | 21 | 47.73 |
| problem list | 122.07 | 339.64 | 15 | 8 | 18.18 |
| notes | 39.32 | 61.80 | 9 | 8 | 18.18 |
| data | 14.44 | 31.32 | 8 | 7 | 15.91 |
| use | 11.92 | 30.83 | 9 | 6 | 13.64 |
| information | 20.20 | 42.28 | 9 | 6 | 13.64 |
| structured | 43.50 | 58.50 | 6 | 6 | 13.64 |
| ward | 40.68 | 53.04 | 6 | 5 | 11.36 |
| system | 10.84 | 17.33 | 5 | 5 | 11.36 |
| note | 17.96 | 24.66 | 5 | 5 | 11.36 |

*Note.* n = 44 responses. Reference corpus: Spoken BNC2014. Range = number of responses containing the keyword.

##### Suggested improvements

Suggestions for improving Epic concerned principally how to use problem lists (29.7% of comments) and templates (18.9%), improved access to important information (10.8%), and the quality and quantity of training (10.8%) (Table 5). Concordance analysis showed that suggested improvements to problem lists and templates centred on greater flexibility to make tailored adjustments, increased integration across parts of the system (for example, making it easier to transfer and access information between problem lists and discharge summaries), and better explanation of both why and how such features should be used. Respondents wanted to understand why entering structured data mattered, not only how to do it, and suggestions about training were focused on these same areas.

**Table 5.** Keywords from corpus analysis of open-text responses on suggested improvements to the EHR.

| Keyword | Simple Maths | Log-likelihood | Frequency | Range | Range % |
| --- | --- | --- | --- | --- | --- |
| use | 29.01 | 82.16 | 16 | 12 | 32.43 |
| problem list(s) | 101.22 | 72.89 | 13 | 11 | 29.73 |
| to use | 54.00 | 72.89 | 10 | 8 | 21.62 |
| how to | 40.85 | 53.06 | 9 | 7 | 18.92 |
| template(s) | 55.77 | 71.26 | 8 | 7 | 18.92 |
| access(ed)(ible) | 25.39 | 28.33 | 7 | 5 | 10.81 |
| training | 28.48 | 34.79 | 6 | 4 | 10.81 |

*Note.* n = 37 responses. Reference corpus: Spoken BNC2014.

### Study 2: IPA findings

Analysis generated four Group Experiential Themes (GETs). An overview of all GETs and their constituent Personal Experiential Themes (PETs) is provided in Table 6.

**Table 6.**
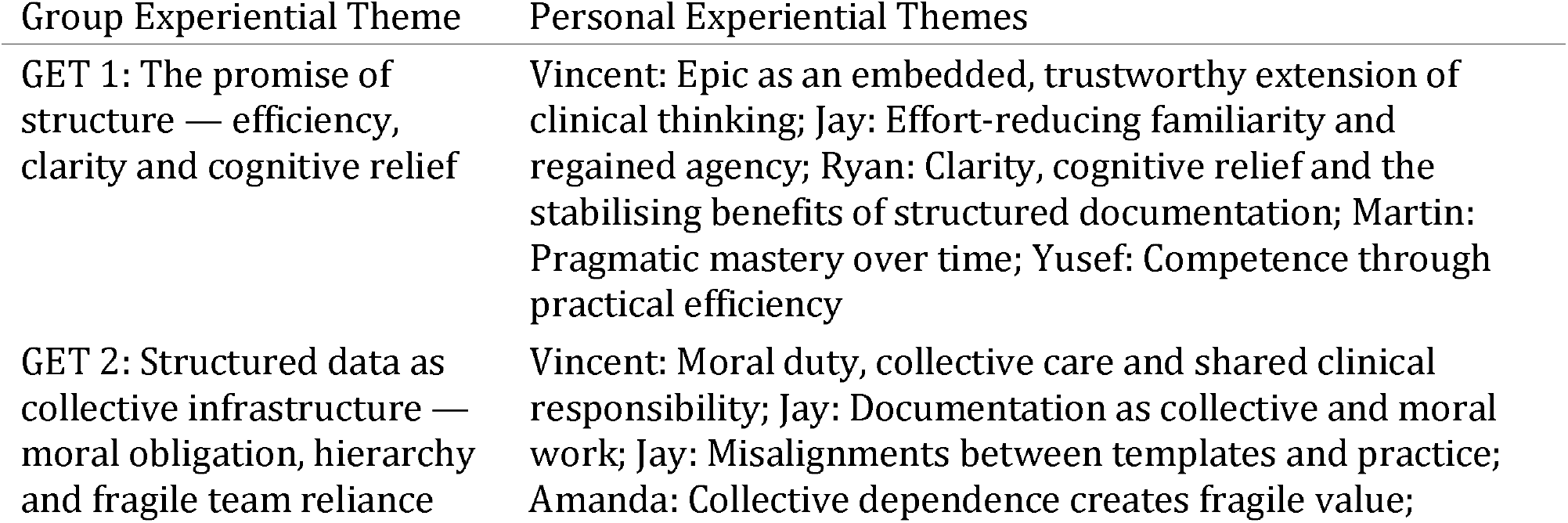
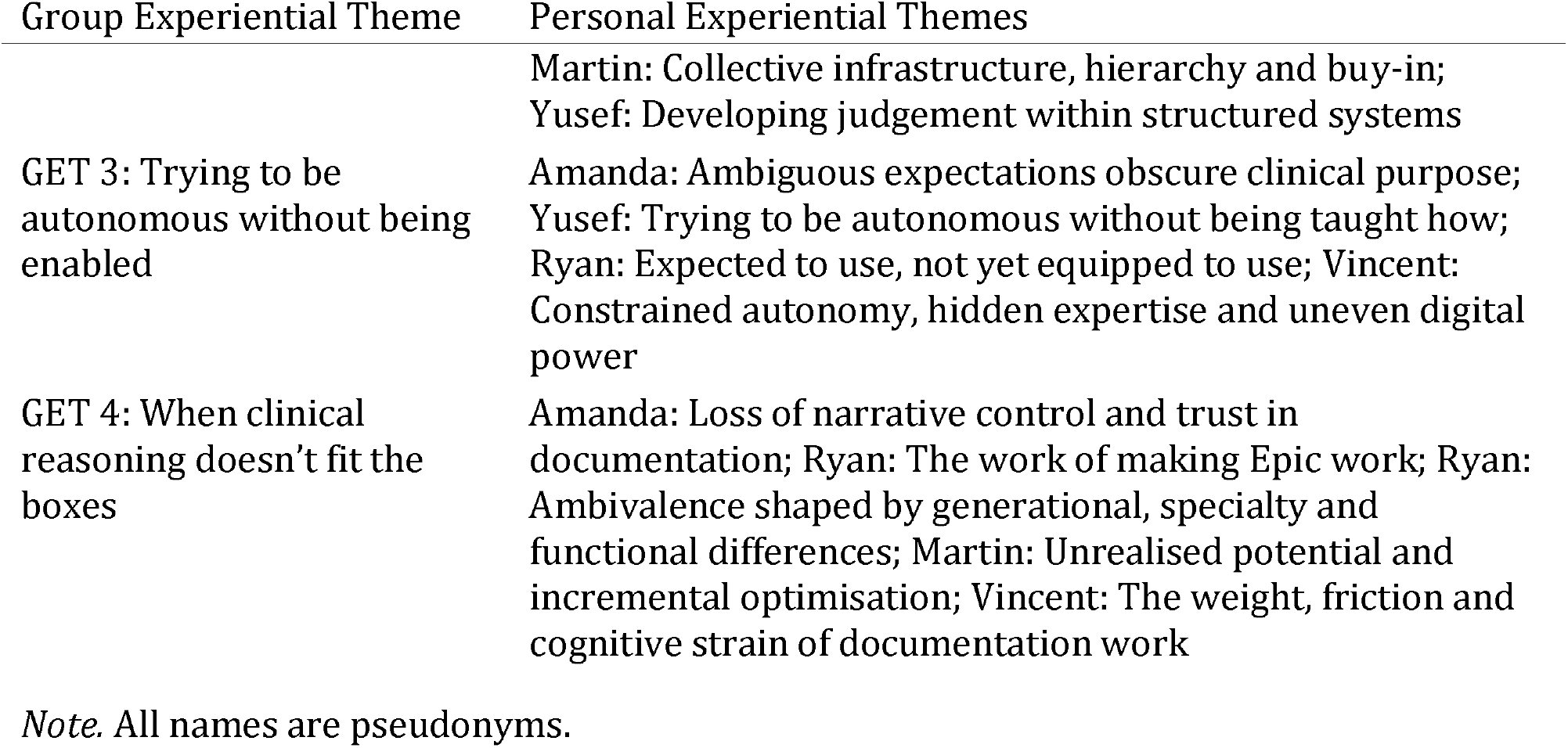
Group Experiential Themes and constituent Personal Experiential Themes. Group Experiential Theme Personal Experiential Themes.

#### GET 1: The promise of structure — efficiency, clarity and cognitive relief

Across the accounts, Epic is not experienced simply as a record system but as something that reshapes how clinical work is mentally carried. When it aligns with practice, it is lived as a form of cognitive support that lightens the burden of remembering, tracking and reconstructing patient information. Doctors describe a shift from holding complexity internally to encountering it as something already organised and visible, producing a sense of stability and reassurance in otherwise demanding clinical environments:

> “Pre-made templates are really helpful definitely. It again saves time […] it also helps you to remind things which you might otherwise forget when you are doing a clinical consultation. […] Problem list […] that’s again very helpful because it’s at a glance, it gives you everything that’s been happening with the patient. Less chances of forgetting. Less chances of missing that.” (Yusef, haematology ST5)

The emphasis here is not only on speed but on a felt reduction in cognitive risk: information is encountered as already ordered and “at a glance”, easing the pressure to remember and mentally reconstruct the patient’s story. This experience gains further meaning in contrast to memories of paper notes, recalled as more fragmented, slower and more effortful:

> “It’s really easy to access any information the moment it’s entered […] which will be quite difficult with paper.” (Vincent, haematology ST2)

The immediacy of access marks a shift in how time and effort are experienced in clinical work. Waiting, searching and piecing together information are replaced by a sense of continuity and availability, reinforcing Epic’s role as a dependable cognitive support. Over time, familiarity deepens this relationship, transforming the system from something merely used into something that can be actively shaped. Customisation becomes a way of aligning the system with one’s own way of working, and with that a sense of regained agency emerges:

> “You can customise your view. So the things that I use most commonly, I can put them on top […] So whatever I want, I have put it on top of my chart […] 80, 90% of the time, whatever you need, it is right there in front of my view.” (Jay, haematology MST7)

Here the experience is one of fluency and fit. Beyond individual use, the structure of the system is also experienced as supporting continuity and shared understanding across the team. Standardised formats and cumulative problem lists allow patient information to be encountered as a developing narrative rather than a series of disconnected entries:

> “They standardise notation so that everyone on different days of the week, no matter who’s documenting, you get a very similar output of structure, of note, and then when it comes to looking back or looking at other ward round notes, when you’re seeing a patient it’s easy to, you’re very familiar […] in-build problem lists allows you to create a progressive, additive note as the problem evolves, develops. You’re easily able to create a picture of what’s been happening.” (Ryan, haematology registrar)

In these moments, Epic becomes a stable reference point that helps doctors situate themselves within the patient’s ongoing and multidisciplinary story. Taken together, these accounts articulate the promise of structure as it is lived in practice: a promise of cognitive relief, visual clarity and psychological reassurance. When the system works well, it is experienced not as an additional task but as a supportive extension of clinical thinking.

#### GET 2: Structured data as collective infrastructure — moral obligation, hierarchy and fragile team reliance

Across interviews, doctors do not experience Epic as a private workspace but as a shared organisational infrastructure. Documentation is understood as a form of collective labour, animated by moral, professional and interpersonal obligations to colleagues who rely on the accuracy and completeness of the record. Entries are made not only for one’s own use but for the benefit of others:

> “You should document as soon as you can, depending on your workloads, for other people to be able to access your information […] your role, reason why we’re seeing patient, all the findings you have, your impression, and the plan […] respond to the messages of your colleagues in proper time to make everyone effective.” (Vincent, haematology ST2)

This shared responsibility is nonetheless experienced as fragile. The effectiveness of the system is contingent on consistent contribution by others, which is often lacking. Doctors repeatedly describe frustration at having to compensate for incomplete notes, outdated problem lists or insufficiently detailed entries left by colleagues. Rotational staffing exacerbates this fragility: norms are inconsistent, expectations unclear, and senior–junior divides shape how documentation is approached:

> “What you can’t have is incomplete buy in […] if you’re going to have a problem list, that has to be maintained. So every single person has to be maintaining it […] if other people aren’t going to use it, it’s going to become outdated very quickly and it becomes kind of useless […] otherwise you’re just wasting your time.” (Amanda, haematology registrar)

Hierarchical differences are especially salient. Junior doctors rely heavily on structured formats for clarity, safety and guidance, while senior clinicians, supported by experience and clinical intuition, use the system more selectively. This asymmetry generates tension: juniors feel pressured to use structured fields precisely because seniors depend on them, yet seniors themselves rarely model or reinforce the behaviours required to keep the system functioning well:

> “The way most of us juniors use EPIC is different to how one of our senior colleagues will. We will tend to stretch it to its absolute limits in search of efficiency […] but the responsibilities of senior colleagues are different […] they are very strict on using the templates because they allow them to get the information they need. Whereas if they’re doing the same thing, they may not need to use the same template, but they do what’s relevant.” (Martin, junior neurosurgical resident)

The result is a sense that the system’s benefits are collectively produced but individually felt, and that the work of keeping it useful falls unevenly on particular people or groups. Without shared commitment, structured documentation risks becoming unreliable, leading some doctors to disengage. This theme captures the EHR as a collaborative ecosystem: powerful when enacted consistently, but precarious when fractured by uneven buy-in.

#### GET 3: Trying to be autonomous without being enabled

Across doctors and specialties, structured documentation is something participants are expected to use well but not taught how to use well. Epic is introduced as an integral part of modern clinical work, yet the learning environment surrounding it is fragmented, rushed or insufficiently scaffolded. Induction training is often brief, overwhelming and quickly overshadowed by clinical pressures, leaving doctors equipped only for surface-level use. As they progress, many discover that they do not know what the system can do, which features matter for their specialty, or what “good use” actually looks like. This creates persistent uncertainty, not about the value of the EHR but about how to enact structured documentation in practice:

> “I use it mainly for kind of literally summarising information for myself and other people […] whereas I do feel like technically we are probably supposed to be using the problem lists especially to help with coding and things like that […] I don’t really know what management expects, but I imagine there is a gap between what we’re actually doing and what was expected.” (Amanda, haematology registrar)

The lack of clear expectations forces doctors into reactive, self-directed learning, relying on trial and error, peer observation or informal support rather than structured pedagogy. With no shared benchmarks for adequacy, they struggle to judge how much detail is expected, when templates should be adapted, or how to balance completeness against efficiency. This ambiguity undermines confidence, particularly for international medical graduates and newly rotating trainees, who feel pressure to appear competent while still unsure about core functions:

> “One of the important bits of structured data that I’m not using because I don’t have enough knowledge and I haven’t used before is the report. I’ve tried to use them, but I don’t get enough information.” (Vincent, haematology ST2)

Doctors repeatedly describe fears about “breaking” the system, deleting information, or failing to meet an invisible standard. These emotional burdens create hesitancy to explore wider functionality, constraining autonomy and reinforcing a pattern in which the system is used only to the extent that feels safe:

> “The home page of EPIC, it’s so dense with icons and stuff that I don’t get time to explore around. What I was thinking was to try to get a remote access and maybe when I am at home, explore around. But then I’m afraid I might click a wrong button and delete some vital information.” (Yusef, haematology ST5)

Responsibility for bridging these gaps falls disproportionately on individual doctors, often encroaching on personal time, producing exhaustion and a sense that they must survive rather than thrive with the system. In this context, autonomy is experienced as expected but not enabled: a form of structural disempowerment that shapes how the EHR is used, understood and lived.

#### GET 4: When clinical reasoning doesn’t fit the boxes

Alongside difficulties with training and expectations, doctors experience the system itself as a source of friction: a platform that is powerful and widely valued, yet frequently misaligned with clinical reasoning, workflow realities and the nuanced demands of patient care. Features that appear logical in principle, such as structured templates, problem lists and coded categories, often feel rigid or cumbersome in practice. Templates may flatten complex patient narratives into fragmented tick-boxes, obscuring the clinical story and requiring doctors to reconstruct meaning across multiple screens:

> “Because it’s now just a form that people fill out […] whatever falls outside of the stuff that’s just presented to you in the form people are no longer recording […] now it’s just whatever the boxes ask you to do people are filling in, however anything that isn’t that is gone […] the useful bits have kind of been missed […] it’s all the kind of symbolic things rather than actually useful.” (Amanda, haematology registrar)

This misalignment produces a distinctive form of cognitive labour: doctors must continually work around the system to make it usable. They double-check entries, compensate for others’ incomplete documentation, and use narrative notes, free-text fields or off-template strategies to restore lost nuance. When templates do not contain the information they need, doctors document in inappropriate sections simply to ensure that vital details are visible:

> “There’s a standardised way of entering into the notes that a patient has a treatment and escalation plan […] but obviously the nature of those kinds of discussions and those decisions is very nuanced and isn’t the way you’re documenting […] that’s probably one of the times where it’s very difficult to use something like a structured note to codify what you’re inputting because a lot of it is kind of long, difficult conversations that you can’t really boil down to predetermined points that are already coded into EPIC […] using that to represent a difficult conversation I’d felt quite kind of disjointed and not representative.” (Ryan, haematology registrar)

Friction also emerges through interface limitations: clunky navigation, cluttered layouts, slow pathways and dense displays that overwhelm rather than streamline thought. Doctors repeatedly describe the system as having enormous potential, yet feeling unfinished or under-optimised. Features such as problem lists, in-system messaging and treatment plans are praised in principle but experienced as unreliable, inconsistently updated or simply unused by colleagues:

> “I feel like it’s [the Epic chat function] being underused […] that’s one way which EPIC is being underutilised in usage. Its message function hasn’t been optimised the best […] you really have to stretch all of its capabilities in order to maximise efficiency and so I would say I think it could be more efficient.” (Martin, junior neurosurgical resident)

Thus, despite widespread commitment to digital working, doctors articulate a lingering ambivalence: the EHR is indispensable but not yet transformative; useful but not fully aligned; efficient in parts but effortful overall. The friction arises not from resistance to digitisation but from the difficulty of fitting a rigid, system-wide structure to the fluidity of clinical reasoning.

## Discussion

This study examined why non-consultant grade doctors at a large NHS trust do or do not enter structured data in the Epic EHR. The survey identified which beliefs were associated with intention to enter structured data, and the interviews explored how doctors experience structured documentation in their daily work.

### Key findings in relation to existing literature

Concerning our first objective, Epic is valued in principle but experienced as falling short in practice. The below-benchmark SUS score is consistent with prior findings in UK emergency departments [49] and with international evidence linking poor EHR usability to reduced productivity, patient safety risk and burnout [15-17]. The relative underuse of structured features mirrors variability documented across US hospital systems [20, 50, 51] and is now demonstrated in a UK context. Crucially, the Study 2 data establish that this underuse does not reflect principled resistance: GET 1 shows that structured documentation, when it works, is experienced as genuine cognitive relief. GET 4 demonstrates how interface rigidity can nonetheless drive reversion to narrative notes even among clinicians otherwise positively disposed to SDE.

Concerning our second objective, the predominance of attitudes and subjective norms over perceived behavioural control in predicting intention aligns with prior TPB research in healthcare, where evaluative and social factors frequently carry greater weight than perceived control [28, 29]. GET 2 illuminates the normative mechanism in particular: the norms driving documentation behaviour are not abstract professional standards but the immediate, observable behaviour of senior colleagues, and when seniors do not model structured entry the normative environment becomes demanding but unsupported. This matters in a rotational training system. Much of the benefit of high-quality structured data, in coding, service planning and secondary analysis, accrues to the institution over a timescale longer than a typical placement, whereas the effort is borne immediately by doctors who will have moved on. The misalignment between who invests and who benefits is a plausible contributor to the attitudinal barriers observed here and should be considered when designing interventions.

On perceived behavioural control, the bivariate correlation with intention (r = 0.57) suggests it is not irrelevant, and two explanations for its failure to predict independently should be considered. The first is statistical, with 49 complete responses, the regression was underpowered to detect small independent contributions, and a non-significant coefficient should not be read as evidence of absence. The second is a measurement problem. The final PBC composite comprised two items, one capturing capacity and one capturing controllability, the other two having been removed for low internal consistency. A two-item measure spanning two conceptually distinct facets of control is a thin basis for a multivariate test, and treating PBC as unidimensional is a recognised limitation of the model as commonly applied [27]. GET 3 makes the consequence concrete: clinicians do not experience low control simply as an inability to perform the behaviour, but as a more fundamental uncertainty about what competent use even looks like, a state no two-item scale could adequately capture. Time pressure was the most common barrier in the open-text responses (47.7%), so respondents were clearly aware of situational constraints. The PBC items may not have separated lack of time from uncertainty about what adequate practice requires. Future work would benefit from a fuller control measure in which self-efficacy, autonomy and situational control are modelled as distinct predictors. It is also worth noting that, in the TPB, PBC is theorised to exert a direct influence on behaviour as well as an indirect influence through intention [26], a pathway this study was not designed to test.

Concerning our third objective, the barrier profile (e.g. time pressure, interface friction, inadequate training, and the relative ease of narrative documentation) closely replicates findings reported using a different framework in a different national context [24], suggesting that these are structural features of any transition from narrative to structured documentation rather than site-specific problems. The IPA data add that training barriers operate at a deeper level than prior literature has captured: GET 3 describes not simply practical incompetence but epistemic uncertainty, with clinicians unsure what adequate structured documentation looks like and afraid to explore features they were never taught. This is consistent with evidence that one-time EHR training rapidly becomes inadequate as systems evolve [11], and with calls for ongoing, specialty-specific support rather than generic induction [34].

Concerning our final objective, the Study 2 themes extend the quantitative findings in two directions. First, GET 2 reveals that structured documentation is experienced as collective moral work done for colleagues and future clinicians rather than for compliance, which implies that individual-level behaviour change interventions will be insufficient without normative change at team level. This aligns with implementation evidence that partial adoption undermines the value of system-wide features and that sustained uptake requires shared commitment not individual motivation alone [52]. Second, GET 4’s account of friction between structured templates and clinical reasoning reflects a tension documented in emerging work on the transition from free text to structured documentation [53], and suggests that the goal should be complementary rather than competing documentation modes, preserving narrative space within structured systems rather than displacing it entirely.

### Implications for intervention design

The number of interventions designed to facilitate effective EHR use has grown in recent years [33, 34]. Interventions focused on improving specific EHR functions and enhancing data quality have shown significant effectiveness, being five times more likely to improve EHR use compared with controls [33]. Our findings point to recommendations in four areas.

#### Training

Education is among the most effective and feasible intervention categories [34]. Both the survey data and the IPA accounts (GET 3) identify training as fragmented, non-specific and insufficiently scaffolded. Interventions should prioritise practical, real-time educational methods including simulated patient cases, live demonstrations and peer mentoring. Training should also cover both the “how” and the “why” of SDE, clarifying what constitutes adequate use, what is expected, and why it matters for patient care, team communication and data quality. Newly rotating trainees may need particular support.

#### User interface redesign

The below-benchmark SUS score and the friction described in GET 4 both point to a need for more intuitive design. Enhancements should focus on reducing the time burden of SDE, for example by simplifying problem list maintenance through automation, natural language processing [54] or ambient documentation technology [55]; by improving template flexibility and specialty customisation; and by ensuring faster navigation [19]. Interface design should also keep room for free text, because participants described templates that left no place for clinically important detail. Department-led, user-centred optimisation processes offer one practical model for achieving this [18, 56].

#### Workflow integration

Organisational interventions that streamline data entry and enable smoother transitions between modules, for example linking problem lists with discharge summaries, are associated with improved uptake [33, 34]. The IPA data suggest that buy-in across teams and grades matters, and that technical solutions not accompanied by normative reinforcement at team level are unlikely to sustain adoption.

#### Social and normative influences

In the survey, attitudes and subjective norms were the strongest correlates of intention. In the interviews, norms appeared as what seniors did and expected, what the team expected, and a sense of obligation to colleagues who relied on the record. Interventions should promote a culture that values SDE, for example by encouraging enthusiastic, experienced peers within clinical teams to provide local support, and by ensuring that organisational leadership reinforces these norms through policy, visible support and performance feedback [34]. Participants described problem lists becoming outdated, and therefore unhelpful, when only part of the team maintained them (GET 2).

### Strengths and limitations

This study has several methodological strengths. In the mixed-methods design, the survey identified which beliefs were associated with intention, and the interviews described how doctors experience those influences in practice, which helps interpret the survey results. The use of the TPB provides a replicable, theory-driven framework that allows direct comparison with prior intervention research, while IPA provides a detailed account of how the practice is experienced. The corpus linguistic analysis offers a systematic, reproducible way to analyse free-text survey responses, and we are not aware of its previous use in EHR research. Finally, the study adds UK NHS evidence to a literature dominated by US research conducted under different organisational and financial conditions.

Several limitations should be noted. The survey sample comprised predominantly resident doctors from a single institution using a single EHR, which limits generalisability. Because recruitment combined Trust-wide mailing lists with in-person teaching events, the number of doctors reached is unknown and no response rate can be reported. Against an estimated eligible workforce of approximately 1,000 non-consultant grade doctors, 83 responses represent a small fraction, and self-selection is therefore a potential concern as doctors holding strong views about the EHR, in either direction, may have been more likely to respond. The findings describe the views of a self-selected group of doctors and should not be read as prevalence estimates for the Trust. In addition, the analyses were powered to detect large effects only, and the non-significant PBC coefficient should be interpreted accordingly. Finally, the cross-sectional design precludes causal inference.

Future research should include more diverse samples across geographical locations, institutional settings, different EHR systems and clinical specialties, with multi-centre or cross-country designs offering broader perspective on the usability and training challenges observed. Qualitative work should sample across a wider range of specialties to examine whether experiences vary systematically by clinical context. Longitudinal studies tracking behaviour following targeted interventions would help establish whether improvements in training and interface design produce enduring change, and objective metrics such as system logs and time-stamped interaction data would strengthen the self-reported behavioural measures used here.

### Conclusions

This mixed-methods study examined the factors associated with structured data entry among non-consultant grade doctors using the Epic EHR at a large NHS Trust. There are four main findings. First, attitudes and subjective norms were independently associated with intention to enter structured data. Perceived behavioural control was not, although this may reflect limited power and a two-item measure. Second, respondents rated Epic’s usability below the SUS benchmark of 68. Third, interviewees described structured documentation as both helpful and limiting. It eased the work of keeping track of patients, but it could not always capture clinical reasoning, depended on the whole team keeping it up to date, and was hard to use well without adequate training. Fourth, the barriers we found involve team norms, training and system design as well as individual beliefs.

Efforts to improve structured data entry are therefore likely to need changes to training, interface design and team norms together, while keeping space for free-text clinical narrative, if structured data are to provide a foundation for high-quality, reusable health data.

## Data Availability

The datasets generated and analysed during the current study are not publicly available. The study was conducted at a single site and the interview transcripts contain contextual detail that could allow individual participants to be identified; participants did not consent to public data sharing, and the terms of ethical approval do not permit it.

## List of abbreviations

EHR: Electronic health record
SDE: Structured data entry
TPB: Theory of Planned Behaviour
PBC: Perceived behavioural control
SUS: System Usability Scale
IPA: Interpretative Phenomenological Analysis
GET: Group Experiential Theme
PET: Personal Experiential Theme
NHS: National Health Service
UCLH: University College London Hospitals NHS Foundation Trust
SAS: Specialist, Associate Specialist and Specialty
COREQ: Consolidated criteria for reporting qualitative research
STROBE: Strengthening the Reporting of Observational Studies in Epidemiology.

## Declarations

### Ethics approval and consent to participate

Ethical approval for both studies was granted by the University College London Research Ethics Committee (approval ID 005). All participants received a participant information sheet and provided informed consent electronically before taking part. Interview participants additionally provided written and verbal consent to audio and video recording. All methods were carried out in accordance with the Declaration of Helsinki and relevant institutional guidelines and regulations.

### Consent for publication

Not applicable. All quotations are anonymised and participants are identified by pseudonym only.

### Availability of data and materials

The datasets generated and analysed during the current study are not publicly available. The study was conducted at a single site and the interview transcripts contain contextual detail that could allow individual participants to be identified; participants did not consent to public data sharing, and the terms of ethical approval do not permit it. Aggregated data may be available from the corresponding author on reasonable request, subject to the conditions of the ethical approval. The survey instrument and interview guide are provided as Additional Files 1 and 2.

### Competing interests

This research was funded by UCLH Charity, the charitable arm of the NHS Foundation Trust at which the study was conducted. The funder had no role in the design of the study, in the collection, analysis or interpretation of data, or in the decision to submit for publication. The authors declare no other competing interests.

#### Funding

This research was funded by UCLH Charity. The study was conducted as an independent piece of research by University College London. The funder had no role in the design of the study, in the collection, analysis or interpretation of data, in the writing of the manuscript, or in the decision to submit it for publication.

#### Authors’ contributions

Author contributions are reported using the CRediT taxonomy. SB: Conceptualization, Methodology, Software, Formal analysis, Investigation, Data curation, Visualization, Project administration, Writing – original draft, Writing – review & editing. CS: Conceptualization, Methodology, Formal analysis, Investigation, Writing – original draft, Writing – review & editing. AR: Conceptualization, Methodology, Writing – review & editing. AG: Conceptualization, Methodology, Supervision, Writing – review & editing. AJW: Conceptualization, Methodology, Resources, Funding acquisition, Writing – review & editing. All authors contributed to the theoretical framing of the study, and all authors read and approved the final manuscript.

## Acknowledgements

The authors thank the doctors who gave their time to complete the survey and to take part in interviews, and colleagues at University College London Hospitals NHS Foundation Trust who supported distribution of the survey and recruitment of interview participants.

## Additional files

Additional File 1. Survey instrument (Theory of Planned Behaviour questionnaire, System Usability Scale as administered, training and feature-use items, open-text prompts). Format: DOCX.

Additional File 2. Semi-structured interview topic guide. Format: DOCX. Additional File 3. STROBE checklist for cross-sectional studies. Format: DOCX. Additional File 4. COREQ checklist. Format: DOCX.

